# No associations between physical activity and clinical outcomes among hospitalized patients with severe COVID-19

**DOI:** 10.1101/2020.11.25.20237925

**Authors:** Ana J. Pinto, Karla F. Goessler, Alan L. Fernandes, Igor H. Murai, Lucas P. Sales, Bruna Z. Reis, Mayara Diniz Santos, Hamilton Roschel, Rosa M. R. Pereira, Bruno Gualano

**Affiliations:** Applied Physiology and Nutrition Research Group, School of Physical Education and Sport; Clinical Hospital HCFMUSP, School of Medicine FMUSP, University of Sao Paulo, Sao Paulo-SP, Brazil; Rheumatology Division, Clinical Hospital HCFMUSP, School of Medicine FMUSP, University of Sao Paulo, Sao Paulo-SP, Brazil; Food Research Center, University of Sao Paulo, Sao Paulo-SP, Brazil

**Keywords:** SARS-CoV-2, prognosis, lifestyle, physical inactivity

## Abstract

**Purpose:** This small-scale, prospective cohort study nested within a randomized controlled trial aimed to investigate the possible associations between physical activity levels and clinical outcomes among hospitalized patients with severe COVID-19.

**Methods:** Hospitalized patients with severe COVID-19 were recruited from Clinical Hospital of the School of Medicine of the University of Sao Paulo (a quaternary referral teaching hospital), and from Ibirapuera Field Hospital, both located in Sao Paulo, Brazil. Physical activity levels were assessed by Baecke Questionnaire of Habitual Physical Activity. The primary outcome was hospital length of stay. The secondary outcomes were: mortality, admission to the intensive care unit (ICU), and mechanical ventilation requirement.

**Results:** Mean hospital length of stay was 8.5 ± 7.1 days; 3.3% of patients died, 13.8% were admitted to ICU, and 8.6% required mechanical ventilation. Linear regression models showed that physical activity indexes were not associated with hospital length of stay (work index: β=-0.57 [95%CI: −1.80 to 0.65], p=0.355; sport index: β=0.43 [95%CI: −0.94 to 1.80], p=0.536; leisure-time index: β=1.18 [95%CI: −0.22 to 2.59], p=0.099; total activity index: β=0.20 [95%CI: −0.48 to 0.87], p=0.563. Physical activity indexes were not associated with mortality, admission to ICU and mechanical ventilation requirement (all p>0.05).

**Conclusions:** Among hospitalized patients with COVID-19, physical activity did not associate with hospital length of stay or any other clinically-relevant outcomes. These findings suggest that previous physical activity levels may not change the prognosis of severe COVID-19.

## Introduction

Physical inactivity has been considered a predisposing factor to acquired infections in several cohorts (1-3). A solid body of literature provides biological plausibility for the negative impact of physical inactivity on the immune system. For instance, chronic inactivity has been linked to increased systemic inflammation, impaired natural killer cell cytolytic activity (4), and reduced T-cell proliferation and cytokine production (5, 6), which can ultimately lead to a loss of viral control (7).

The immunoregulatory role of physical activity is also well known. Regular physical activity has been postulated to improve, or at least maintain, immunity across the life span (8). Evidence has also pointed out that physical activity is able to reduce the incidence and the number and severity of symptoms associated with acute respiratory infections (e.g., upper respiratory tract infection) (9).

The link between physical (in)activity and COVID-19 remains to be properly stablished. A recent study showed that physical inactivity increases the relative risk for COVID-19 hospital admission in a UK cohort by 32% (10), suggesting that the adoption of an active lifestyle could lower the risk of severe infection. Nonetheless, it is unclear whether physical activity confers a better prognosis to patients hospitalized with severe COVID-19.

This small-scale, prospective cohort study aimed to investigate the possible associations between physical activity levels and clinical outcomes (hospital length of stay, mortality, intensive care unit (ICU) admission and mechanical ventilator requirement) among hospitalized patients with severe COVID-19.

## Methods

### Study design

This is a prospective, cohort study nested within a multicenter, randomized controlled trial designed to test the safety and efficacy of vitamin D_3_ supplementation in hospitalized patients with severe COVID-19 (ClinicalTrials.gov Identifier: NCT04449718), conducted between June 2, 2020 and October 7, 2020. We assessed patients’ clinical status, coexisting chronic diseases, demographic characteristics, self-reported body weight and height, ethnicity, and physical activity upon hospital admission. Clinical outcomes were assessed through medical records.

### Patients

Hospitalized patients were recruited from the Clinical Hospital of the School of Medicine of the University of Sao Paulo (a quaternary referral teaching hospital), and from Ibirapuera Field Hospital, both located in Sao Paulo, Brazil. Inclusion criteria were: 1) adults aged 18 years or older; 2) diagnosis of COVID-19 by either polymerase chain reaction (PCR) for severe acute respiratory syndrome coronavirus 2 (SARS-CoV-2) from nasopharyngeal swabs or computed tomography scan findings (bilateral multifocal ground-glass opacities ≥ 50%) compatible with the disease; 3) diagnosis of flu syndrome with hospitalization criteria on hospital admission, presenting with respiratory rate ≥ 24 breaths per minute, saturation < 93% on room air or risk factors for complications, such as heart disease, diabetes mellitus, systemic arterial hypertension, neoplasms, immunosuppression, pulmonary tuberculosis, and obesity, followed by COVID-19 confirmation. Exclusion criteria were: 1) patient unable to read and sign the written informed consent; 2) patient already admitted under invasive mechanical ventilation; 3) previous vitamin D_3_ supplementation (> 1000 IU/day); 4) renal failure requiring dialysis or creatinine ≥ 2.0 mg/dL; 5) hypercalcemia defined by total calcium > 10.5 mg/dL; 6) pregnant or lactating women; and 7) patients with expected hospital discharge in less than 24 hours upon admission.

The study was approved by the Ethics Committee of Clinical Hospital of the School of Medicine of the University of Sao Paulo and by the Ethics Committee of Ibirapuera Field Hospital. All the procedures were conducted in accordance with the Declaration of Helsinki. The participants provided written informed consent before being enrolled in the study (approval number: 30959620.4.0000.0068).

### Outcome measures

Physical activity levels were assessed by Baecke Questionnaire of Habitual Physical Activity (11), which has been previously validated to the Brazilian population (12). The questionnaire consists of 3 sections: work, sport, and leisure-time activity. Scores in each section range between 0 and 5, where higher scores indicate a higher physical activity level. A total activity index is obtained by summing all scores (maximum score = 15).

The primary outcome was hospital length of stay, defined as the total number of days that patients remained hospitalized from the date of hospital admission until the date of hospital discharge or death. The criteria used for patient discharge were: 1) no need for supplemental oxygen in the last 48 hours; 2) no fever in the last 72 hours; and 3) oxygen saturation > 93% in room air without respiratory distress. The secondary outcomes were: 1) mortality; 2) admission to the ICU; and 3) mechanical ventilation requirement.

### Statistical Analysis

Associations between physical activity levels (independent variables) and hospital length of stay (primary outcome) were tested using linear regression models, whereas associations between physical activity levels and mortality, admission to ICU, and mechanical ventilation requirement (secondary outcomes) were tested using logistic regression models. In a sensitivity analysis for the primary outcome, patients were divided into tertiles according to total activity index. Then, potential differences between tertiles were assessed by generalized estimating equations (GEE) model, based on the assumption of a normal distribution, identity link function, and an exchangeable working correlation, with group (lower, mid, and upper tertiles) as fixed factor. All regression and GEE models were adjusted for potential confounders (i.e., age, sex, BMI, and number of comorbidities). For the primary outcome, models were also adjusted for mortality. Statistical analyses were performed in SAS, version 9.4. Data are expressed as β and 95% confidence interval (95%CI) or odds ratio (OR) and 95%CI. Significance level was set at p=0.050.

## Results

### Patients

Two-hundred and nine patients had complete physical activity data and were included in this study. Patients’ age was 54.9 ± 14.5 years, BMI was 31.0 ± 6.5 kg/m^2^, 51.7% were men, 48.3% were white, 48.8% had hypertension, 28.7% had diabetes, and 11.5% had cardiovascular diseases (Table 1). Seventy-three percent required supplemental oxygen (134 were on oxygen therapy and 19 were on non-invasive ventilation), and 62.2% had computed tomography scan findings suggestive of COVID-19. Mean hospital length of stay was 8.5 ± 7.1 days, 3.3% of patients died, 13.8% were admitted to ICU, and 8.6% required mechanical ventilation. Mean work index was 2.1 ± 0.8 (range: 0.9 to 3.8), sport index was 2.0 ± 0.7 (range: 1.0 to 4.3), leisure-time index was 2.5 ± 0.7 (range: 1.0 to 4.3), and total activity index was 6.6 ± 1.5 (range: 3.1 to 11.1).

**Table 1.** Demographic and clinical characteristics.

|  | n=209 |
| --- | --- |
| Age, mean (SD), y | 54.9 (14.5) |
| Sex, No. (%) |  |
| Male | 108 (51.7) |
| Female | 101 (48.3) |
| Race, No. (%) |  |
| White | 101 (48.3) |
| Brown | 78 (37.3) |
| Black | 30 (14.4) |
| Body mass index, mean (SD), kg/m<sup>2</sup> [n=192] | 31.0 (6.5) |
| Underweight, No. (%) | 2 (1.0) |
| Normal, No. (%) | 28 (14.6) |
| Overweight, No. (%) | 58 (30.2) |
| Obesity, No. /total (%) | 104 (54.2) |
| Acute COVID-19 symptoms, No. (%) |  |
| Fever | 149 (71.3) |
| Cough | 177 (84.7) |
| Fatigue | 184 (88.0) |
| Arthralgia | 71 (34.0) |
| Myalgia | 144 (68.9) |
| Nasal congestion | 78 (37.3) |
| Coryza | 87 (41.6) |
| Sore throat | 69 (33.0) |
| Diarrhea | 95 (45.5) |
| Coexisting diseases, No. (%) |  |
| Hypertension | 102 (48.8) |
| Cardiovascular disease | 24 (11.5) |
| Diabetes | 60 (28.7) |
| Chronic obstructive pulmonary disease | 7 (3.3) |
| Asthma | 10 (4.8) |
| Rheumatic disease | 17 (8.1) |
| Concomitant medications, No. (%) |  |
| Antibiotic | 207 (99.0) |
| Anticoagulant | 188 (90.0) |
| Analgesic | 139 (66.5) |
| Corticosteroids | 156 (74.6) |
| Antihypertensive | 101 (48.3) |
| Hypoglycemic | 42 (20.1) |
| Hypolipidemic | 28 (13.4) |
| Antiemetic | 117 (56.0) |
| Antiviral | 3 (1.4) |
| Proton pump inhibitor | 112 (53.6) |
| Levothyroxine | 17 (8.1) |
| Physical activity level, mean (SD) |  |
| Work index | 2.1 (0.8) |
| Sport index | 2.0 (0.7) |
| Leisure-time index | 2.5 (0.7) |
| Total activity index | 6.6 (1.5) |

3 Data are expressed as mean (SD) or absolute and relative frequencies (n[%]).

### Primary Outcome

Linear regression models showed that physical activity indexes were not associated with hospital length of stay (work index: β=-0.50 [95%CI: −1.60 to 0.60], p=0.368; sport index: β=0.09 [95%CI: −1.14 to 1.32], p=0.8823; leisure-time index: β=0.90 [95%CI: −0.37 to 2.17], p=0.162; total activity index: β=0.07 [95%CI: −0.53 to 0.68], p=0.810; Figure 1A).

**Figure 1.**
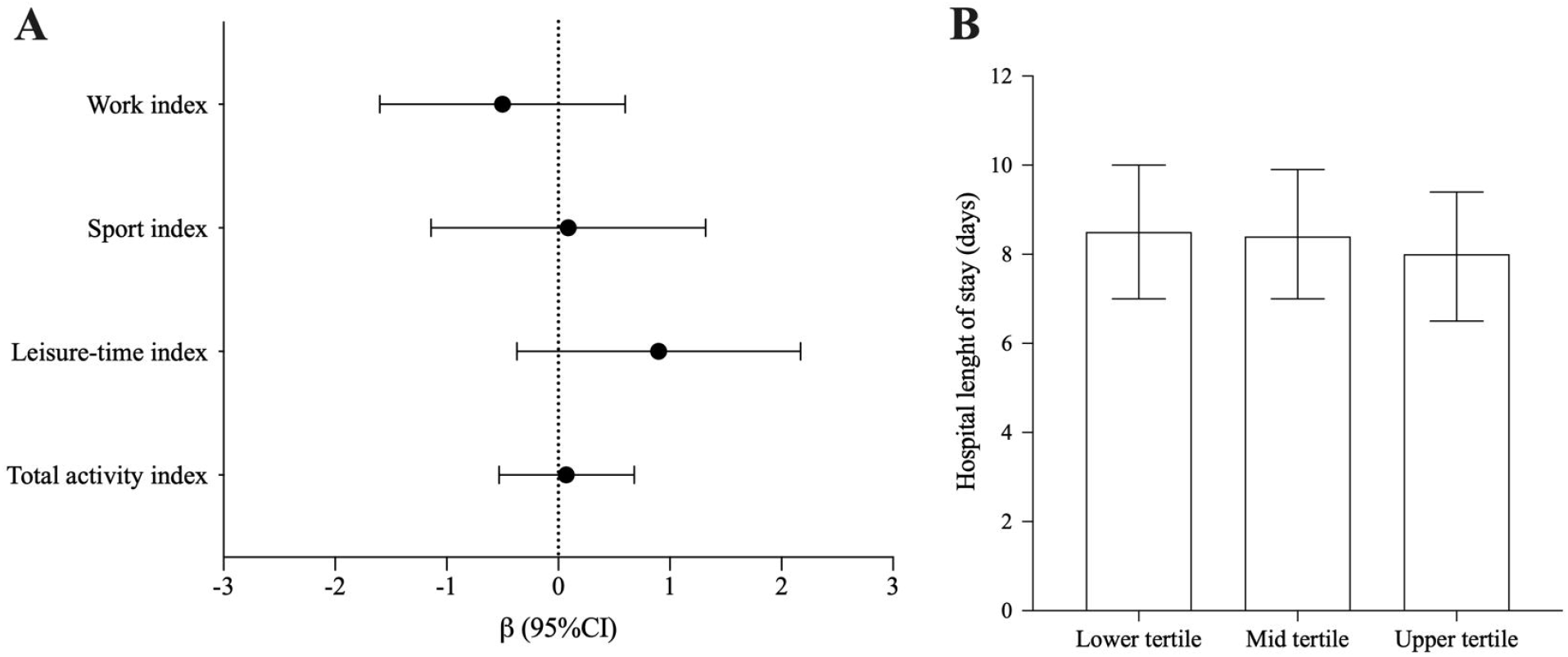
Associations between physical activity level and hospital length of stay. Data are presented as β (95%CI).

Lower (5.0 [95%CI: 4.9 to 5.2]), mid (6.6 [95%CI: 6.5 to 6.8]), and upper (8.3 [95%CI: 8.1 to 8.4]) tertiles significantly differed in respect of total activity index (p<0.001). Sensitivity analysis showed that hospital length of stay was comparable between lower (8.5 days [95%CI: 7.0 to 10.0]), mid (8.4 days [95%CI: 7.0 to 9.9]) and upper (8.0 days [95%CI: 6.5 to 9.4]) tertiles of total activity index (p=0.875; Figure 1B).

### Secondary Outcomes

Logistic regression models showed that physical activity indexes were not associated with mortality (work index: OR=1.1 [95%CI: 0.3 to 3.8], p=0.936; sport index: OR=0.4 [95%CI: 0.1 to 1.5], p=0.192; leisure-time index: OR=0.5 [95%CI: 0.1 to 2.1], p=0.342; total activity index: OR=0.7 [95%CI: 0.4 to 1.3], p=0.272), admission to ICU (work index: OR=-1.1 [95%CI: 0.6 to 2.0], p=0.697; sport index: OR=0.9 [95%CI: 0.5 to 1.8], p=0.867; leisure-time index: OR=0.5 [95%CI: 0.3 to 1.1], p=0.077; total activity index: OR=0.9 [95%CI: 0.7 to 1.2], p=0.459), and mechanical ventilation requirement (work index: OR=0.6 [95%CI: 0.3 to 1.2], p=0.176; sport index: OR=0.9 [95%CI: 0.4 to 2.0], p=0.844; leisure-time index: OR=0.7 [95%CI: 0.3 to 1.6], p=0.413; total activity index: OR=0.8 [95%CI: 0.5 to 1.2], p=0.214; Figure 2).

**Figure 2.**
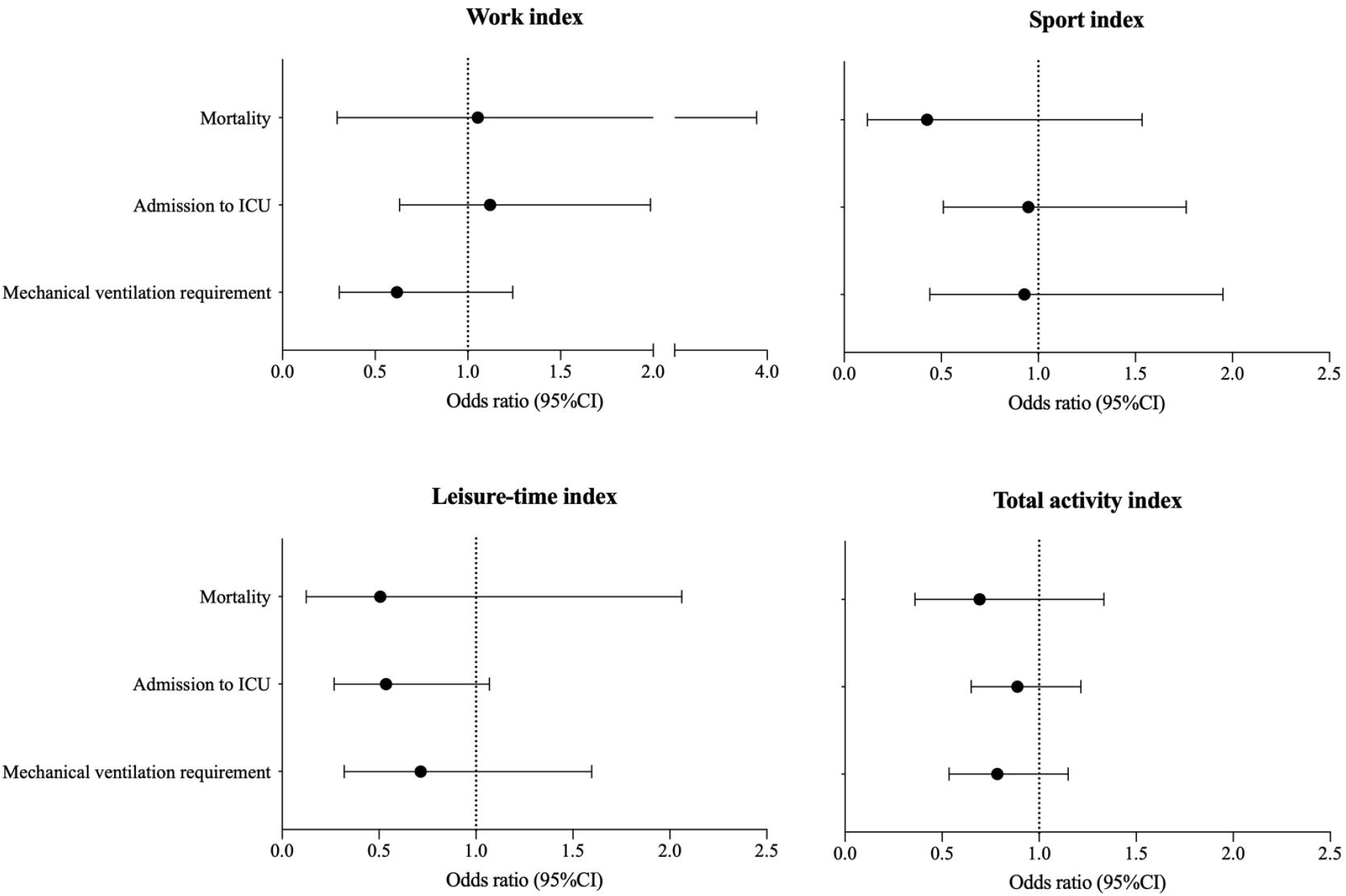
Associations between physical activity level and mortality, admission to ICU, and mechanical ventilation requirement. Data are presented as OR (95%CI).

## Discussion

This small-scale, prospective, cohort study provided novel evidence that physical activity levels do not associate with hospital length of stay or any other relevant clinical outcomes among patients with severe COVID-19.

The negative impact of physical inactivity on the immune system has been widely reported (4-7). Inactivity has been associated with higher rates of viral and bacterial infections, immunosenescence, and poor vaccine responses (4-7). In addition, inactivity is closely related to obesity, a condition that predisposes to poor antibody responses to vaccination (13, 14) and impaired lymphocyte proliferation following mitogenic stimulus (15). These immunological disturbances, among others, experienced by obese people are implied in a greater risk of viral and bacterial infections as well as longer hospital length of stay due to more frequent and prolonged complications following surgery (16, 17). This body of literature has supported the speculation that physical inactivity could be a risk factor for COVID-19 as well.

A few recent studies have supported this hypothesis. A large-scale, observational study involving individuals residing in England showed that physical inactivity and obesity account for up to 8.6% and 29.5%, respectively, of the population attributable fraction of severe COVID-19 (10), defined as a case requiring hospital admission. These findings are corroborated by another study showing that being physical active is associated with a 35% reduction in hospitalization due to COVID-19 in Brazil (18). While these studies suggest that physical activity may prevent hospital admission due to COVID-19, it remained to be investigated the influence of lifestyle on the prognosis in already hospitalized patients.

This study aimed to address this gap by following a small cohort of hospitalized patients with severe COVID-19. We showed that physical activity levels did not associate with hospital length of stay or any clinical outcome. It is difficult to contrast these findings with others since there is a lack of analogous studies involving COVID-19 patients. However, among patients with cardiovascular disease, higher physical activity level was related to a slightly shorter hospital stay (0.9 day) (19). A similar conclusion was reached by another study that observed that physical inactivity was a predictor of longer hospital lengths of stay among community-dwelling older adults (3.18 vs 0.82 days in active individuals) (20). One may speculate that the main factor underlying the contrasting outcomes found in these cohorts and in ours are the patients’ characteristics. In fact, the longer hospital length of stay in the current study (8.5 days vs 2.6 (20) and 2.1 days (19)) appears to indicate that our patients were more severe than those of the other cohorts. Furthermore, it is possible to conjecture that the protective effect of physical activity may be less pronounced in acute infectious diseases, such as COVID-19, as compared to chronic conditions, such as cardiovascular diseases (19).

The main strength of this study involves its novelty in assessing the influence of physical activity levels, using a validated questionnaire, in a hospitalized cohort of patients with severe COVID-19 and several comorbidities (see Table 1).

The main limitations of this study involve its observational nature, which hampers causative relationships; the small sample size, which increases the chances of type 2 errors, particularly considering the low incidence for the secondary outcomes; the use of a questionnaire to assess physical activity, which is prone to recall bias, overreporting, and limited time frame of evaluation (i.e., 1 year); and the lack of patients’ follow-up after the hospital discharge.

In conclusion, among hospitalized patients with COVID-19, physical activity did not associate with hospital length of stay or any other clinically relevant outcomes. These findings, which needs further replication in larger cohorts, suggest that previous physical activity levels may not change the prognosis of severe COVID-19.

## Data Availability

The datasets used and/or analyzed during the current study are available from the corresponding author on reasonable request.

## Acknowledgments

A.J.P., K.F.G., I.H.M., A.L.F., R.M.R.P., and B.G. were supported by São Paulo Research Foundation – FAPESP (grants #2015/26937-4, #2019/18039-7, #2019/24782-4; #2020/11102-2, #2016/00006-7 and #2020/05752-4, and #2017/13552-2). R.M.R.P., H.R., and B.G. were supported by Conselho Nacional de Desenvolvimento Científico e Tecnológico (grants 305556/2017-7, 301571/2017-1, and 305242/2019-9).

## Conflict of interest

The authors declare no conflict of interests. Additionally, authors declare that results of the study are presented clearly, honestly, and without fabrication, falsification, or inappropriate data manipulation. Finally, the results of the present study do not constitute endorsement by ACSM.

## Notes

### Competing Interest Statement

The authors have declared no competing interest.

### Funding Statement

A.J.P., K.F.G., I.H.M., A.L.F., R.M.R.P., and B.G. were supported by Sao Paulo Research Foundation - FAPESP (grants #2015/26937-4, #2019/18039-7, #2019/24782-4; #2020/11102-2, #2016/00006-7 and #2020/05752-4, and #2017/13552-2). R.M.R.P., H.R., and B.G. were supported by Conselho Nacional de Desenvolvimento Cientifico e Tecnologico (grants 305556/2017-7, 301571/2017-1, and 305242/2019-9).

### Author Declarations

The study was approved by the Ethics Committee of Clinical Hospital of the School of Medicine of the University of Sao Paulo and by the Ethics Committee of Ibirapuera Field Hospital (approval number: 30959620.4.0000.0068).

